# Registered Report: Artifact Index for Capacitive Electrocardiography Acquired with an Armchair

**DOI:** 10.64898/2026.06.03.26353526

**Authors:** Joana M Warnecke, Diana Baumgärtel, Julian Bollmann, Thomas M Deserno

## Abstract

1

**Background:** Continuous health monitoring enables early detection of diseases and improves therapeutic outcomes. Non-intrusive biosignal sensors, such as capacitive ECG (cECG), offer a practical solution for daily monitoring in private environments, such as smart homes and vehicles. However, artifacts reduce signal quality and compromise reliability.

**Methods:** Following a registered report protocol (Warnecke JM et al. Plos One. 2021; 16(7):e0254780), we record data of 44 subjects and develop an artifact index for cECG. We use three signal quality indices (SQIs): the correlation of QRS complexes (corSQI), the R-peak detection consistency (bSQI) and the absolute amplitude ratio (aSQI). Our index classifies overlapping 10 s segments with a step-width of 2 s into clean or artifact segments. We label a 2 s interval as artifacts if all five overlapping segments indicate artifacts. We record cECGs using an armchair with integrated electrodes in a single-arm study involving 44 subjects performing two activities — reading and watching television (TV); for 11 minutes each. We record a time-synchronized reference ECG with skin electrodes on the chest. To evaluate the artifact index, we compare it with manually generated ground truth. Moreover, we evaluate the clothing materials cotton, linen, jeans, and polyester in 5 subjects.

**Results:** Watching TV results in longer, continuously clean signal durations than reading. On average, 88.3% of the signal has a minimum continuous clean duration of 10 s, versus 79.8% during reading. All clothing configurations achieve a clean signal duration exceeding 10 s. Among the SQI metrics, bSQI performs best, achieving an accuracy of 90.7% and an F1 score of 79.9%. Combining the three SQI metrics in a voting approach improves accuracy to 92.0% and F1 score to 82.1%.

**Discussion:** Our artifact index automatically distinguishes clean from artifact cECG segments, promoting health monitoring in unsupervised real-world settings, earlier disease detection, and preventive health management. A limitation is the investigation of only two scenarios (reading and watching TV).

## 2 Introduction

New technologies for personalized preventive health monitoring, particularly for continuous monitoring in private spaces, such as the smart home [1] or the smart vehicle [2], support early disease detection and improve therapeutic outcomes [3].

A smart armchair with integrated biosignal recording supports unobtrusive health monitoring in daily routines. The time people spend on activities such as watching television (TV), reading or relaxing makes the armchair an ideal setting for continuous health monitoring. If equipped with sensors, such as electrocardiogram (ECG), it seamlessly monitors biosignals while seated [4], but clean and accurate data is essential.

Hence, data quality is challenging, as various factors affect the capacitive ECG (cECG) signals. For instance, movements, electromagnetic disturbances, fabric wrinkles, ground connection, and coupling capacitance degrade the signal quality [5] with more noise and artifacts as compared to galvanic recording [6].

Sirtoli et al. differ the approaches to reduce noise: material-based, analog circuits, and digital signal processing [7]. For instance, hardware adjustments improve the input impedance of the analogue front-end and reduce interference [8]. However, these techniques cannot eliminate all artifacts.

Intensive research focuses on artifact detection and signal quality indices (SQIs) in cECG to improve the reliability and accuracy of ECG. In our registered report protocol, we present related literature [9]. Since the publication of the protocol in 2021, further research has advanced the field.

Škorić report a precision (not accuracy) of 98% – 100% in removing artifacts from cECG signals using quantified signal fluctuations around a linear trend [10]. Kuetche et al. [11] comprehensively review SQIs of four types of metrics: statistical (e.g., skewness, kurtosis), frequency-based (e.g., regulatity), nonlinear (e.g., entropy [12] or fuzzy entropy [13]), and QRS-based (e.g., template matching). They conclude that the most suitable approach for signal quality detection depends on the type of noise or artifact.

Recently, deep learning complements the metric-based approaches. Jin et al. use long short-term memory (LSTM) networks to assess ECG quality [14]. Rahman et al. compare recursive neural networks (RNNs) and convolutional neural networks (CNNs) for noise detection and propose knowledge-based ECG filtering [15]. Furthermore, subject movement from video helps to detect artifacts [16], but requires additional modalities and synchronization. Although these studies make significant contributions to artifact detection in ECG signals, detecting and evaluating the quality of cECG data remains crucial.

In a preliminary study [17], we analyzed SQI metrics designed for galvanic ECG for their usability with cECG. According to our registered report protocol [9], we now address the following research questions:

1. Are cECG data from an armchair suitable for heart rate (HR) and heart rate variability (HRV) calculation?
2. Can an artifact index identify clean and artifact cECG segments?
3. How does clothing affect the quality of cECG?

## 3 Materials and Methods

In this section, we briefly describe the study design (for details, see [9]), introduce the algorithmic approaches for artifact detection, and explain their evaluation.

### 3.1 Sensors

We record a chair-integrated cECG (prototype, Capical GmbH, Braunschweig, Germany; Fig. 1), a reference ECG (BiosignalPlux Explorer, Plux Wireless Biosignals, Lisboa, Portugal), and photopletysmography (PPG) (BiosignalPlux Explorer, Plux Wireless Biosignals, Lisboa, Portugal). We use optical time synchronization (BiosignalPlux Explorer, Plux Wireless Biosignals, Lisboa, Portugal).

**Fig 1.**
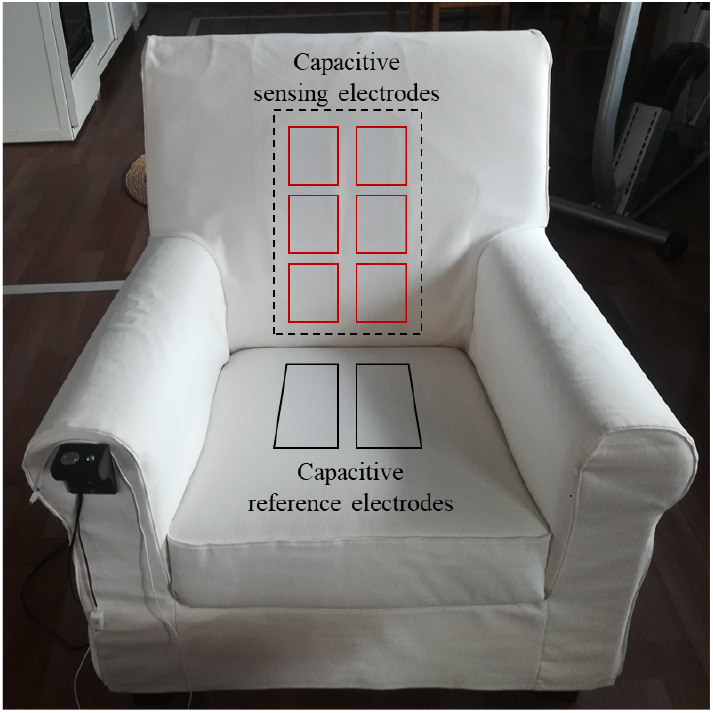
cECG armchair with electrodes [9]

### 3.2 Study Design

We conduct a single-arm study involving 44 subjects and record their anamnestic parameters (i.e., age, gender, size, weight, health status). We have approvals from the data security officer at TU Braunschweig (No: PscECG1) and the ethics committee of Hannover Medical School (No: 9287 BO S2020) for the data security plan and the study protocol, respectively. The recruitment period starts on 01/10/2020 and the end date is 30/09/2021.

The subjects read a book and watch TV for 11 minutes each, while we monitor their biosignals. To minimize any interference with the recorded signals, all subjects wear cotton t-shirts. Additionally, we record 5 subjects to compare the effect of cotton, linen, jeans, and polyester clothes. We do not repeat consistent artifact measurements to ensure a realistic assessment of quality.

### 3.3 Preprocessing

Using vendor software, we calculate 28 cECG channels from the 8 electrodes in the armchair and run a 15-30Hz bandpass filter. We select channel 15 due to its best coverage. We dismiss the first and last 30 s of each recording, leaving 10 minutes for analysis, which we cut into overlapping 10 s segments in steps of 2 s.

### 3.4 Ground Truth

A qualified expert carefully reviews and annotates the full 10 minutes signal, marking periods that contain artifacts (Fig. 2), in particular high-amplitude movement artifacts and equipment noise, omitting low-amplitude artifacts between R-peaks, which are irrelevant for later R-peak detection. If any part of a 2 s segment contains artifacts, we classify the entire segment as such.

**Fig 2.**
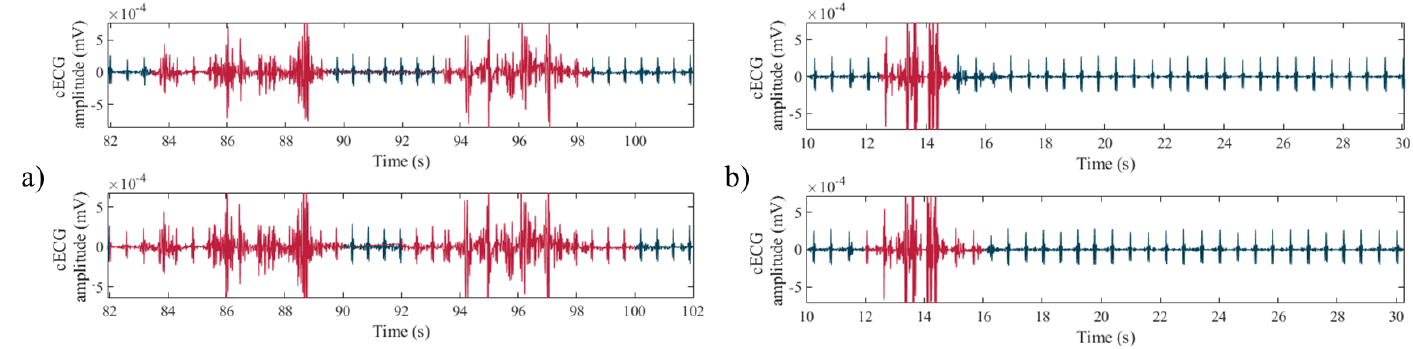
Ground truth annotations before (top) and after (bottom) extension to 2-second segments. The red parts are artifacts. Examples from subject 7 ((a) reading, (b) watching TV).

### 3.5 HR and HRV Measures

Health professionals use 10 s as the standard duration of a clinical 12-lead ECG for HR estimation [18]. We therefore require 10 s ECG recordings without artifacts for HR measurements.

Some relevant metrics for HRV are the standard deviation of normal-to-normal (NN) intervals (StdNN), the square root of the mean squared differences between successive NN intervals (RMSSD), or the number of successive NN interval pairs that differ more than 50 ms (NN50) [19]. Clinicians use long-term (24 h) or short-term (5 min) recordings to calculate the metrics, while researchers investigate ultra-short ECG recordings. Despite a study that shows the validity of 10 s, 30 s, and 120 s for the analysis of RMSSD and 120 s for SDNN [20], Pecchia et al. demonstrate a lack of evidence that ultra-short HRV features surrogate short-term ones [19]. A study by Shaffer et al. shows the sufficiency of 60 s recordings for RMSSD and SDNN for resting individuals [21]. We therefore analyze 10 s, 60 s, 120 s and 300 s segments for HRV measurements.

### 3.6 Artifact Index

Based on a previous review [17], we select three SQIs:

1. *corSQI:* Orphanidou et al. [22] proposed a template-matching signal quality index that quantifies the similarity between individual QRS complexes and an average QRS template. The algorithm first detects R-peaks using the ep limited method [23]. For each beat *i*, it extracts a QRS segment *Q*_*i*_ ∈ ℝ^*L*^ of length *L* around the R-peak and adapts *L* according to neighboring RR intervals. It then constructs a QRS template 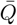 of the average QRS complex of the 10 s segment. For each beat *i*, the algorithm defines the correlation-based SQI as the correlation coefficient between *Q*_*i*_ and the template 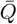:

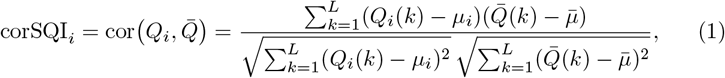

where *µ*_*i*_ and 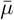 denote the means of *Q*_*i*_ and 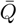, respectively, and *k* = 1, …, *L* indexes the samples within the QRS segment. The algorithm classifies a segment as an artifact if

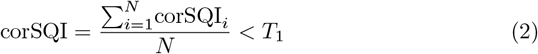

where *N* is the number of peaks and *T*_1_ denotes an empirically chosen threshold.
2. *bSQI*: According to Nayan et al. [24], we determine the ratio of R-peak positions consistently detected with two R-peak detectors

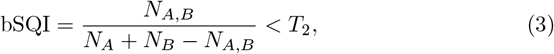

where *N*_*A,B*_ is the number of beats in which both algorithms agree, and *N*_*A*_ and *N*_*B*_ are the number of R-peaks the individual detectors find [11]. As the original implementation is unsuitable for cECG, we use the ep limited [23] and the WQRSM implementation of the R-beat detection algorithm by Moody et al. [25] from the Physionet toolbox [26].
3. *aSQI*: We propose the analysis of amplitudes [17]. We partition the 10 s segments of cECG(*t*) into 5 parts of 2 s and calculate the ratio of the lowest and the highest maximum absolute values. The values are the maximum amplitudes in the parts

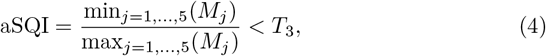

where 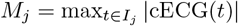 and *I*_*j*_ = [2(*j* − 1), 2*j*]. Values above and below a threshold *T*_3_ indicate uniformity and the presence of differing amplitudes (i.e., artifact), respectively.

Due to the unbalanced class distribution, we optimize the thresholds *T*_*i*_ by F1-score and not by accuracy. According to Orphanidou et al. [22], we determine all SQIs values and define 100 equidistant thresholds between the minimum and maximum values, and a brute force algorithm finds the best.

We combine the SQIs in a majority voting approach, such that two or more must agree on an artifact within the 10 s segment (Fig. 3). In other words, if two (or more) of the SQIs indicate a clean 10 s segment; we classify the segment as such. We optimize all three thresholds together with the brute force algorithm described above. Using our sliding-window approach, we flag the 2 s parts of the signal as artifacts if all of the 5 overlapping 10 s segments are artifact (Fig. 4).

**Fig 3.**
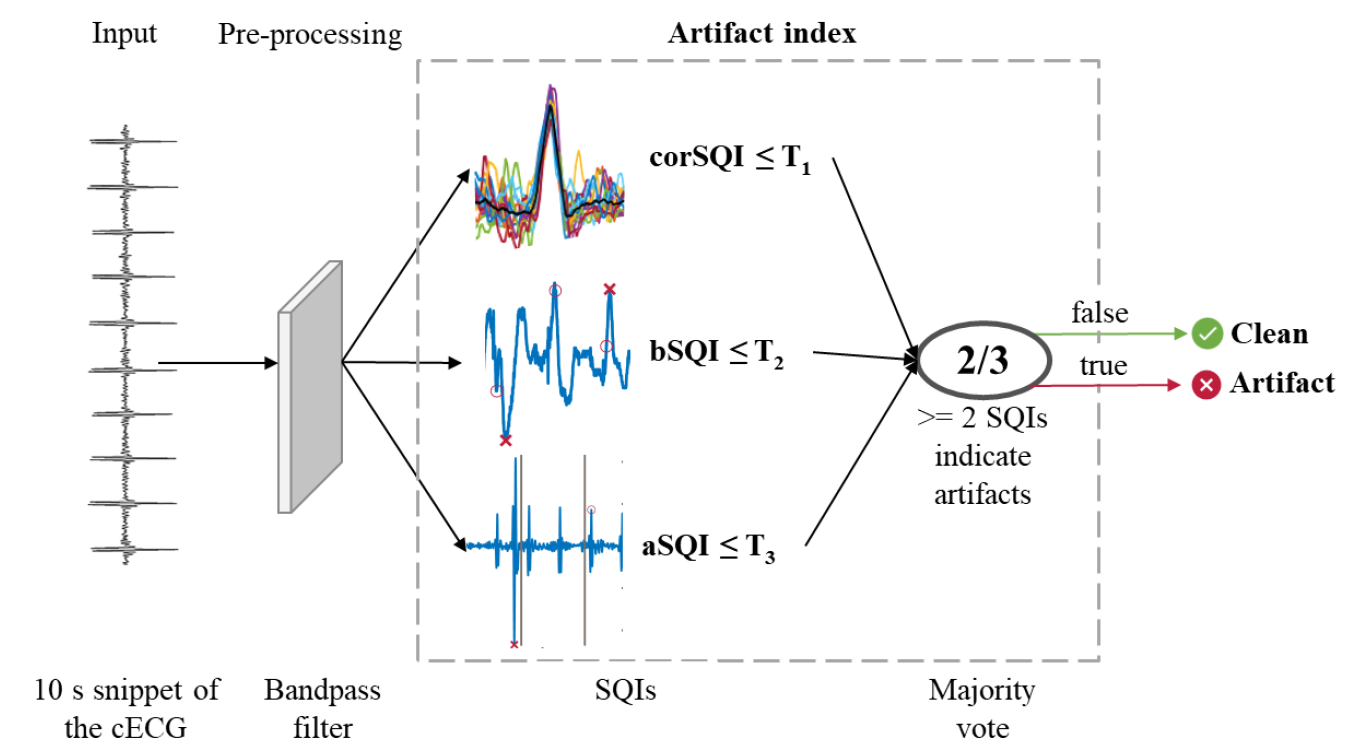
Data pipeline for 10 s segment artifact index.

**Fig 4.**
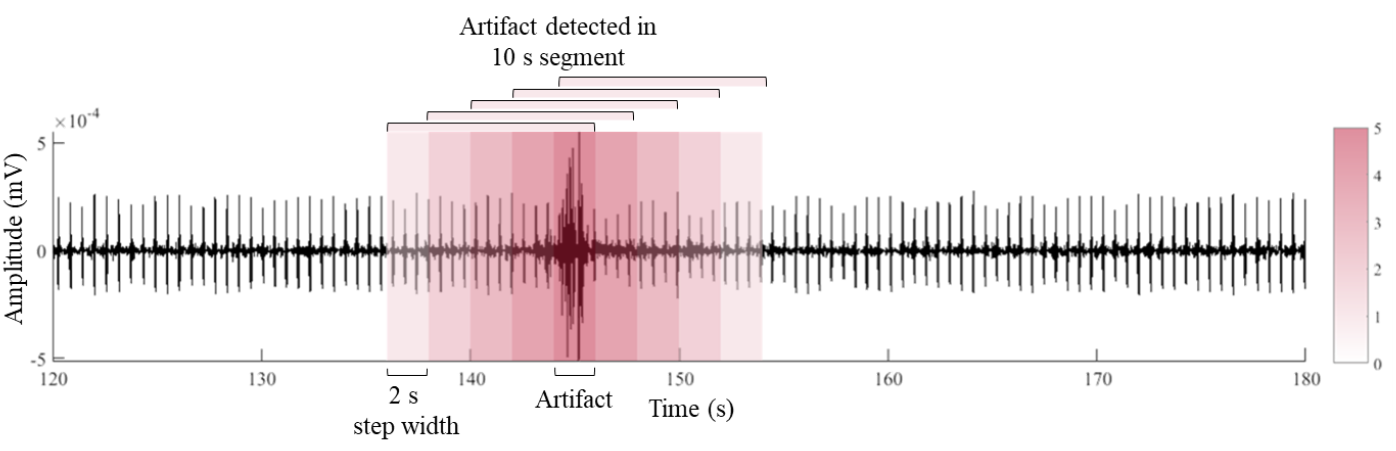
Reliability classification with sliding windows. Only dark red indicates artifacts.

### 3.7 Evaluation

We assess the performance of the SQIs using all 10 s segments of all 44 subjects compared to the ground truth (clean vs. artifact). We determine the accuracy, sensitivity, specificity, and F1-score and visualize violin plots.

In addition, we compare the labeled 2 s steps (clean vs. artifact) with the ground truth. Again, we determine accuracy, sensitivity, specificity, and F1 score, and we visualize the confusion matrix.

## 4 Results

### 4.1 Data

Following the registered report protocol [9], we include 44 subjects, 43 having a clean health status and 1 reporting a non-cardiovascular disease that does not affect the ECG(Tab. 1). Due to a recording error, two of the signals in the jeans scenario are only 9 and 8 minutes, yielding in 8 and 7 minutes of data, respectively.

**Table 1.** Anamnestic parameters of our dataset (44 subjects according to [9]).

| Parameter | Median (Range) | Frequency |
| --- | --- | --- |
| Age | 27 (18-56) years |  |
| Gender |  | 24 male, 20 female |
| Height | 175 (147-195) cm |  |
| Weight | 74 (46-140) kg |  |
| BMI | 23.9 (15.3-43.7) |  |
| Health status |  | 43 clean, 1 non-relevant disease |

### 4.2 Signal Quality

For reading and watching TV, five and two subjects, respectively, deliver only artifacts. The remaining subjects deliver segments of at least 10 s clean signal. Reading and watching TV yield, on average, 79.8% and 88.3% of the recording with at least 10 s of clean duration and 22.7% and 44.6% have a continuous clean duration of 300 s, respectively (Tab. 2). Four (9.1%) and 10 (22.7%) subjects have complete 10 min recordings of clean duration while reading and watching TV, respectively (Fig. 5).

**Fig 5.**
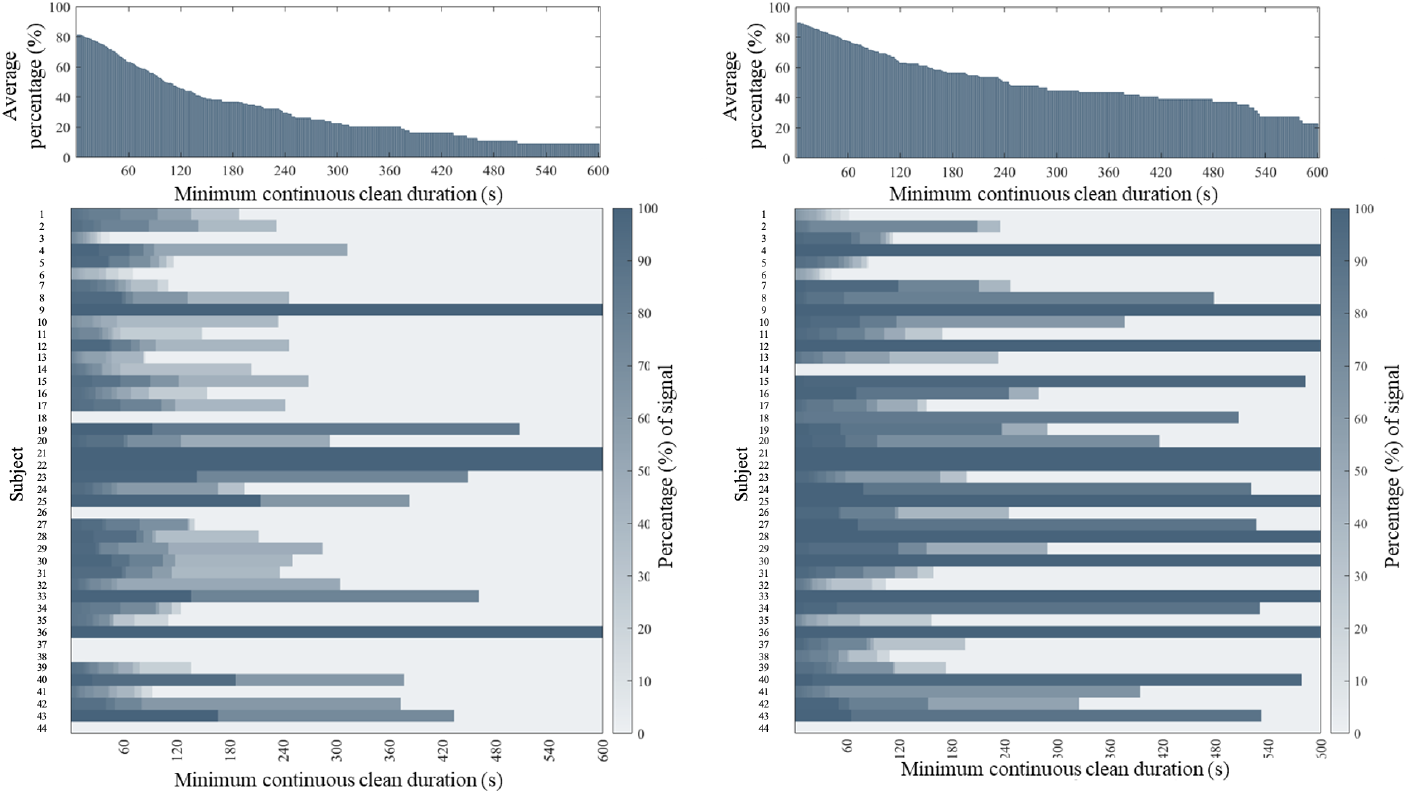
Clean segments (in %) by minimum total duration (top, histogram) and by subject (bottom, heatmap) for the activities reading (left) and watching TV (right)

**Table 2.** Average continuous clean signal durations of each activity.

| Activity | 10 s | 60 s | 120 s | 300 s |
| --- | --- | --- | --- | --- |
| Reading | 79.8% | 63.0% | 45.3% | 22.7% |
| TV | **88.3%** | **77.1%** | **62.6%** | **44.6%** |

Across different clothing materials, all cECG signals have a clean signal duration of over 10 s, sufficient for HR calculation. 17.8% of the polyester-scenario signals have a continuous clean duration of at least 300 s, 15.7% of the cotton-scenario, 10.5% of the jeans-scenario, and 7.7% of the linen scenario (Tab. 3). Again, the TV scenario yields longer clean signal durations (Fig. 6).

**Fig 6.**
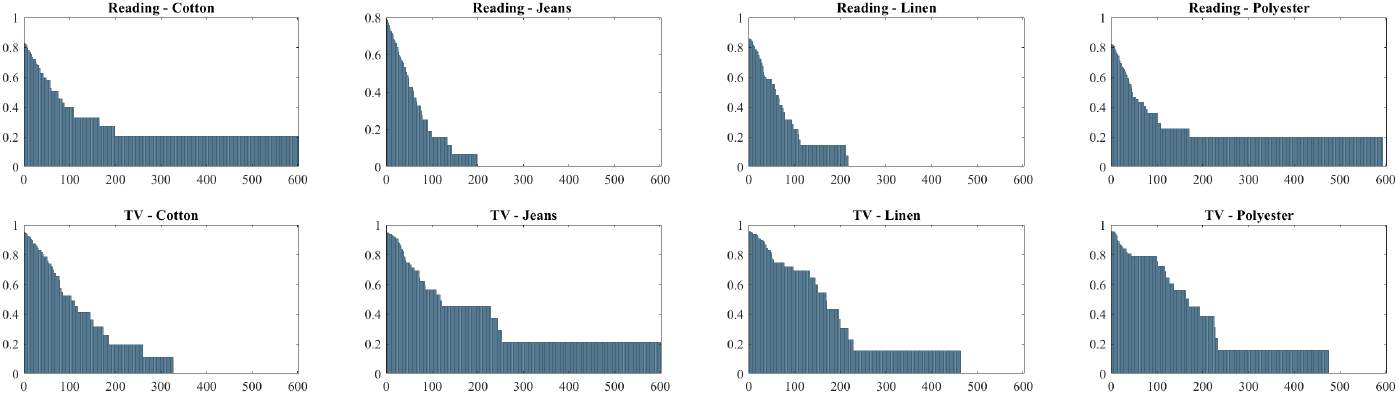
Percentage of clean segments by duration for each activity and clothing material.

**Table 3.** Average continuous clean signal durations per clothing material.

| Clothing material | 10 s | 60 s | 120 s | 300 s |
| --- | --- | --- | --- | --- |
| Cotton | 85.2% | 61.1% | 36.9% | 15.7% |
| Jeans | 83.1% | 54.0% | 32.4% | 10.5% |
| Linen | **87.7%** | **61.2%** | 41.5% | 7.7% |
| Polyester | 85.8% | 61.0% | **45.0%** | **17.8%** |

### 4.3 SQI Performance

Method bSQI performs best (accuracy: 90.7%, F1-score: 79.9%), followed by aSQI (accuracy: 90.0%, F1-score: 75.7%) and corSQI (accuracy: 89.4%, F1-score: 77.7%) (Tab. 4).

**Table 4.** Performance of SQIs for 10 s segments.

| Method | $T_1$ | $T_2$ | $T_3$ | Accuracy | Sensitivity | Specificity | F1-score |
| --- | --- | --- | --- | --- | --- | --- | --- |
| corSQI | 0.8024 |  |  | 89.4% | 85.0% | 90.6% | 77.7% |
| bSQI |  | 0.8788 |  | 90.7% | 85.5% | 92.1% | 79.9% |
| aSQI |  |  | 0.7653 | 90.0% | 71.8% | 95.0% | 75.7% |
| Our SQI | 0.6750 | 0.9793 | 0.7879 | **92.0%** | 85.2% | 94.8% | **82.1%** |

The distribution of SQI values is similar between reading and watching TV (Fig. 7). The value range of bSQI and aSQI is [0,1], the value range of corSQI is [-1, 1]. Our SQI (majority voting) yields best accuracy and F1-score, 92.0% and 82.1%, respectively.

**Fig 7.**
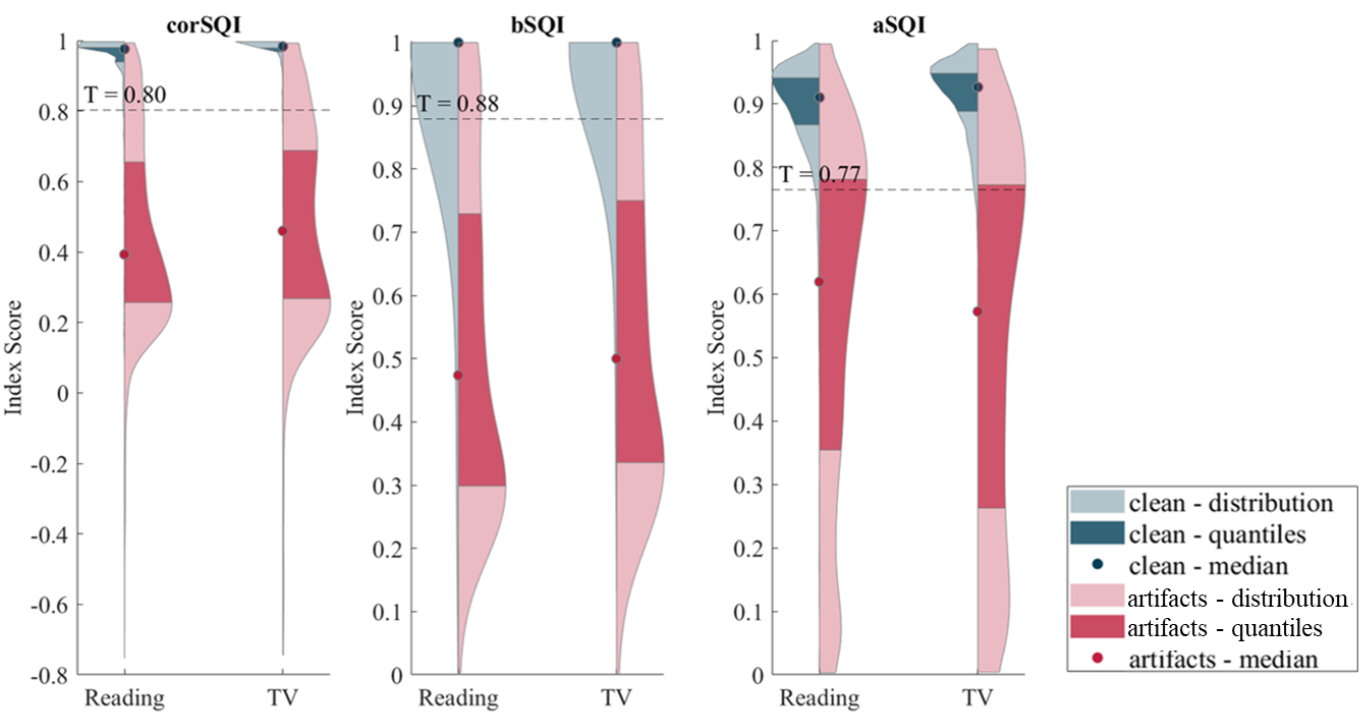
Index value distributions for reading and watching TV, showing thresholds, comparing clean and artifact segments.

### 4.4 Artifact Index Performance

The comparison of expert labels to artifact index results on 2 s segments yields 4.4% FP and 2.3% FN (Tab. 5). The artifact index has an accuracy, sensitivity, specificity, and F1-score of 93.4%, 73.9%, 97.3%, and 78.9%, respectively.

**Table 5.** Confusion matrix for 2 s segments.

|  | True artifact | True clean |  |
| --- | --- | --- | --- |
| Detected artifact | 12.4% | 4.4% | 16.7% |
| Detected clean | 2.3% | 81.0% | 83.3% |
|  | 14.6% | 85.4% | 100% |

## 5 Discussion

Recording with our armchair, 79.8% and 88.3% of the cECG signal with a minimum length of 10 s is clean for reading and watching TV, respectively, but only 22.7% and 44.6% is clean for continuous 300 s, respectively. Across all durations, watching TV yields a higher percentage of clean signal. We explain this with the subjects moving more during reading, which creates artifacts. This shows that the armchair recordings are suitable for HR detection, but signal stability remains a challenge for longer recordings needed for HRV calculation, especially in more movement-prone situations such as reading.

Using a sliding window, our artifact index can identify cECG intervals with a duration of 2 s. It classifies the signal quality of 2 s and 10 s segments with an accuracy of 93.4% and 92.0%, respectively. Only 4.4% of 2 s segments are falsely detected as artifact, and only 2.3% are falsely considered as clean. Therefore, our artifact index appropriately identifies clean and artifact cECG segments in 2 s time steps.

The influence of clothing materials on cECG quality is lower than the influence of the different activities. The availability of 10 s segments (usable for HR detection) ranges between 83.1% and 87.7% of the total signal on average for the different materials. The availability of 300 s segments (usable for HRV detection) lies between 7.7% and 17.8%. Polyester clothing achieves the highest percentage of longer signals, contradicting previous findings [27, 28]. Because we base these clothing-related observations on only five subjects, we recommend further studies. Possibly, clothing thickness, fabric wrinkles, equipment noise, or subtle subject movements have influenced the signal quality more than the material itself [5, 6].

Comparing the individual SQIs shows that all three classify clean and artifact segments with similar, relatively high accuracy, around 90%. This confirms earlier findings by Castro et al., who report accuracies of 77%-91% (depending on the lead) for both bSQI and corSQI on cECG data [29]. Kuetche et al. evaluate different SQIs on galvanic ECGs using the area under ROC curve (AUC), and report results between 89% and 95% AUC for bSQI and 91% to 99% AUC for corSQI for different artifact sources [11]. In both studies, corSQI outperforms bSQI; in our study, bSQI performs slightly better, which is plausible given the different settings.

The majority vote approach improves both accuracy and the F1-score by combining the strengths of the SQIs and, presumably, increasing robustness across different noise types. This aligns with our previous findings [17].

In addition to the registered report protocol [9], we analyze signal durations to gain a deeper understanding of signal quality for medical use. Clinicians typically rely on recordings of a particular length for HR and HRV determination [18, 19], rather than a predefined quality. Consequently, the duration of clean signal segments is crucial for medical analysis and provides more meaningful results than classification.

Although there is still a lack of evidence that ultra-short HRV metrics can replace short-term ones [19, 30], some studies indicate that this the case [20, 21]. Researchers also investigate the possibility of using the average of several very short signal segments (e.g., 10 s) for measurement, improving results from single measurements [20]. If these approaches prove to be valid, significantly larger percentages of the signals are available for measurement: at 60 s instead of 300 s, 63.0% of the signals are usable on average instead of 22.7% for reading and 77.1% instead of 44.6% for TV.

A limitations of our study is that we only perform two activities (reading and watching TV). However, other activities such as playing games or eating will certainly result in even more movement and hence, lower signal quality. Furthermore, our cECG armchair is a prototype, which limits the generalizability of our findings.

Another limitation results from all study participants being free from cardiac diseases, so our findings do not extrapolate to populations with, e.g., arrhythmia. Unlike for galvanic ECG [31, 32], no arrhythmia reference database currently exists for cECG, but extending our algorithm requires such data.

In future research, we will consider data-driven machine learning approaches, e.g., that of Jiang et al., who achieve an accuracy of 99.1% with a selection of SQIs using XGBoost and Shapley additive explanation on their bed-based cECG system [33]. However, the results are not directly comparable with ours as their study setup differs and their dataset is not publicly available.

Moving forward, researchers can use the artifact index to identify clean cECG recordings and exclude artifacts from further analysis. While we analyze continuous health monitoring in an armchair, other settings, such as vehicles [4], offices, and nursing homes are important, and our artifact index serves as a foundation here. With this, we support continuous and personalized health monitoring for preventive medicine.

## 6 Conclusion

- cECG armchair recordings are susceptible to artifacts, necessitating a precise artifact index.
- Signal quality of cECG recordings from an armchair supports HR determination.
- HRV analysis requiring clean recordings of 5 minute length is not possible with the armchair, yet, but approaches to using ultra-short ECG recordings for HRV analysis are being investigated - if they are valid, significantly more of the signal is usable for HRV analysis.
- We contribute a feature-based artifact index for cECG recordings and a freely-available dataset with reference ECG and cECG.

## Data Availability

Public repository: https://leopard.tu-braunschweig.de/receive/dbbs_mods_00079604

## Data Availability

As individual identities cannot be inferred from the ECG and cECG signals, the dataset was published anonymously via the TU Braunschweig Library under a CC BY 4.0 license (https://leopard.tu-braunschweig.de/receive/dbbs_mods_00079604, accessed on December 21st, 2025). The dataset includes reference ECG data (txt format) and cECG data (gen4 format). All participants provided written informed consent and agreed to the publication of these anonymized data.

## Code Availability

The code used for data processing and artifact index computation is publicly available on GitHub (https://github.com/DfromM/artifact-index-cECG, accessed on December 21st, 2025). The repository includes all scripts required to reproduce the analyses presented in this work.

## Acknowledgments

We thank Martin Oehler from Capical for the technical support. We want to extend our gratitude to all the volunteers who participated in our study. Additionally, we deeply appreciate everyone involved in the data recording process.

## References

1. Wang J, Spicher N, Warnecke JM, Haghi M, Schwartze J, Deserno TM. Unobtrusive health monitoring in private spaces: the smart home. Sensors (Basel). 2021 3;28(21):864.

2. Wang J, Warnecke JM, Haghi M, Deserno TM. Unobtrusive health monitoring in private spaces: the smart vehicle. Sensors (Basel). 2021;21(3):864.

3. Steinhubl SR, Waalen J, Edwards AM, Ariniello LM, Mehta RR, Ebner GS, et al. Effect of a home-based wearable continuous ECG monitoring patch on detection of undiagnosed atrial fibrillation: the mSToPS randomized clinical trial. JAMA. 2018;320(2):146–55.

4. Walter M, Eilebrecht B, Wartzek T, Leonhardt S. The smart car seat: Personalized monitoring of vital signs in automotive applications. Pers Ubiquit Comput. 2011;15:707–15.

5. Hou Z, Xiang J, Dong Y, Xue X, Xiong H, Yang B. Capturing electrocardiogram signals from chairs by multiple capacitively coupled unipolar electrodes. Sensors (Basel). 2018;18(9):2835.

6. Wartzek T, Lammersen T, Eilebrecht B, Walter M, S L. Triboelectricity in capacitive biopotential measurements. IEEE Trans Biomed Eng. 2011;58(5):1268–77.

7. Sirtoli VG, Liamini M, Lins LT, Lessard-Tremblay M, Cowan GE, Zednik RJ, et al. Removal of motion artifacts in capacitive electrocardiogram acquisition: a review. IEEE Trans Biomed Circuits and Systems. 2023;17(3):394–412.

8. Xiao Z, Xing Y, Yang C, Li J, Liu C. Non-Contact Electrocardiograms acquisition method based on capacitive coupling. IEEE Instrum Measurem Mag. 2022;25(2):53–61.

9. Warnecke JM, Wang J, Cakir T, Spicher N, Ganapathy N, Deserno TM. Registered report protocol: Developing an artifact index for capacitive electrocardiography signals acquired with an armchair. PLoS ONE. 2021;16(7):e0254780.

10. Škorić T. Reduction of Artifacts in Capacitive Electrocardiogram signals of driving subjects. Entropy. 2022;24(1):13.

11. Kuetche F, Alexendre N, Pascal NE, Colince W, Thierry S. Signal quality indices evaluation for robust ECG signal quality assessment systems. Biomed Physics Eng Expr. 2023;9(5):055016.

12. Fu F, Xiang W, An Y, Liu B, Chen X, Zhu S, et al. Comparison of machine learning algorithms for the quality assessment of wearable ECG signals via lenovo H3 devices. J Med and Biol Eng. 2021;41:231–40.

13. Liu C, Li K, Zhao L, Liu F, Zheng D, Liu C, et al. Analysis of heart rate variability using fuzzy measure entropy. Comp Bio Med. 2013;43(2):100–8.

14. Jin Y, Li Z, Qin C, Liu J, Liu Y, Zhao L, et al. A novel attentional deep neural network-based assessment method for ECG quality. Biomed Sig Proc Control. 2023;79:104064.

15. Rahman S, Pal S, Yearwood J, Karmakar C. Robustness of Deep Learning models in electrocardiogram noise detection and classification. Comp Method Progr Biomed. 2024;253:108249.

16. Srivastava S, Ershadi A, Haghi M, Deserno TM. Reliability estimation of armchair-based capacitive ECG using video-based pose estimation. Conf Proc SPIE Med Imaging. 2023;12469:78–87.

17. Baumgärtel D. Entwicklung eines Artefakt-Index für die Bestimmung der Signalqualität von kapazitiven EKG-Daten. Technische Universität Braunschweig;; 2020. Braunschweig (Germany).

18. Sattar Y, Chhabra L. Electrocardiogram; 2024. Retrieved 2025-12-05. In: StatPearls. Treasure Island (FL): StatPearls Publishing; Jan-. PMID:31747210. Available from: https://www.ncbi.nlm.nih.gov/books/NBK549803/.

19. Pecchia L, Castaldo R, Montesinos L, Melillo P. Are ultra-short heart rate variability features good surrogates of short-term ones? State-of-the-art review and recommendations. Healthcare technology letters. 2018 June;5(3):94–100.

20. Munoz ML, Van Roon A, Riese H, Thio C, Oostenbroek E, Westrik I, et al. Validity of (ultra-) short recordings for heart rate variability measurements. PloS one. 2015;10(9):e0138921.

21. Shaffer F, Shearman S, Meehan ZM. The promise of ultra-short-term (UST) heart rate variability measurements. Biofeedback. 2016;44(4):229–33.

22. Orphanidou C, Bonnici T, Charlton P, Clifton D, Vallance D, Tarassenko L. Signal-quality indices for the electrocardiogram and photoplethysmogram: derivation and applications to wireless monitoring. IEEE J Biomed Health Infor. 2015;19:832–8.

23. Hamilton P. Open source ECG analysis. Conf Proc IEEE Comp Cardio. 2002:101–4.

24. Nayan NA, Risman NS, Jaafar R. A portable respiratory rate estimation system with a passive single-lead electrocardiogram acquisition module. J Europ Soc Eng Med. 2016;24:591–7.

25. Zong W, Moody GB, Jiang D. A robust open-source algorithm to detect onset and duration of QRS complexes. Computers in Cardiology. 2003;30:737–40.

26. Vest AN, Da Poian G, Li Q, Liu C, Nemati S, Shah AJ, et al. An open source benchmarked toolbox for cardiovascular waveform and interval analysis. Physiological Measurement. 2018;39:105004.

27. Hoffmann KP, Ruff R, Poppendieck W. Long-term characterization of electrode materials for surface electrodes in biopotential recording. Conf Proc IEEE Eng Med Biol Soc. 2006;2006:2239–42.

28. Pani D, Achilli A, Bonfiglio A. Survey on textile electrode technologies for electrocardiographic (ECG) monitoring, from metal wires to polymers. J Advanced Materials Technol. 2018;3(10):1800008.

29. Castro ID, Varon C, Torfs T, Van Huffel S, Puers R, Van Hoof C. Evaluation of a multichannel non-contact ECG system and signal quality algorithms for sleep apnea detection and monitoring. Sensors (Basel). 2018;18(2):577.

30. Shaffer F, Meehan ZM, Zerr CL. A critical review of ultra-short-term heart rate variability norms research. Frontiers in neuroscience. 2020;14:594880.

31. Moody GB, Mark RG. The impact of the MIT-HIB arrythmia database. IEEE Eng in Med and Biol. 2001;20(3):45–50.

32. Zheng J, Guo H, Chu H. A large scale 12-lead electrocardiogram database for arrhythmia study. PhysioNet. 2022.

33. Jiang Y, Xiao Z, Zhang Y, Ma C, Yang C, Jin W, et al. An optimized signal quality assessment method for noncontact capacitive ECG. IEEE Trans Instrumen Measurem. 2025;4001711(74):1–11.

